# Effect of a Saffron Extract on Sleep Quality in Adults with Moderate Insomnia: A Decentralized, Randomized, Double-blind, Placebo-Controlled Trial

**DOI:** 10.1101/2025.05.07.25326999

**Authors:** Julius Schuster, Christin Mundhenke, Hannah Nordsieck, Camille Pouchieu, Line Pourtau, Andreas Hahn

**Author notes:** Both authors contributed equally (Julius Schuster, Christin Mundhenke). **Corresponding author**: Julius Schuster, Institute of Food and One Health, Leibniz University Hannover Am kleinen Felde 30, 30167 Hannover, Germany.

## Abstract

**Aim:** Natural interventions for sleep disturbances, such as saffron extract, are gaining scientific and clinical interest. This 3-arm, randomized, double-blind, placebo-controlled trial examined the effect of a standardized saffron extract (Safr’Inside^TM^) on sleep, stress, and other associated psychological outcomes in 165 men and women reporting moderate insomnia.

**Methods:** Participants received a daily capsule containing 30 mg saffron extract, 20 mg saffron extract, or placebo (maltodextrin) for 4 weeks. The primary endpoint was the change in insomnia symptoms measured by the Athens Insomnia Scale (AIS). Secondary outcomes included the Single-Item Sleep Quality Scale (SQS), Perceived Stress Scale (PSS), Patient Health Questionnaire-4 (PHQ-4), Positive and Negative Affect Schedule (PANAS), Epworth Sleepiness Scale (ESS), and World Health Organization Quality of Life (WHOQOL). Analyses followed an intention-to-treat (ITT) approach, with per-protocol (PP) confirmation.

**Results:** Compared to placebo, saffron extract significantly improved insomnia symptoms (AIS, *P* < .05), with the most pronounced effects on sleep onset and duration. Sleep quality (SQS) improved significantly after 3 weeks (*P* < .05 for 30 mg, *P* < .01 for 20 mg). Both doses of saffron extract reduced perceived stress (PSS) more than placebo (*P* = .01). Psychological symptoms (PHQ-4) improved significantly with 30 mg saffron compared to placebo (*P* < .05), while all other measures showed no significant differences. No serious adverse events were reported.

**Conclusions:** Four weeks of 20 or 30 mg saffron extract may reduce insomnia and stress in middle-aged adults. Future research should assess longer interventions, reliable objective sleep measures, and complementary effects of saffron on stress and sleep.

## 1. Introduction

The prevalence of sleep disturbances is increasing worldwide, with approximately 10–45% of the population reporting sleep problems [1]. Certain groups, such as women, older adults, and individuals under chronic stress, are particularly vulnerable to developing sleep disorders [2]. While short-term sleep deprivation can often be compensated by subsequent adequate rest, persistent sleep issues have severe consequences, including impaired physical and mental health, reduced cognitive performance, and lower quality of life [1]. Moreover, sleep disturbances are associated with a 1.74-fold higher risk of cardiovascular disease, while depression increases the likelihood of sleep disturbances 4.07-fold, underscoring the role of sleep disturbance in linking mental and physical health [3]. Conversely, depression itself is known to exacerbate sleep disturbances, creating a cyclical relationship [4].

Conventional treatment approaches for insomnia often begin with non-pharmacological strategies, such as improving sleep hygiene and cognitive behavioral therapy for insomnia (CBT-I) [5]. In more severe cases, pharmacological therapies, including benzodiazepines and non-benzodiazepine receptor agonists, are administered [5]. However, these medications often come with significant side effects, a potential for dependence, and reduced long-term effectiveness [4].

Given these limitations, alternative therapies have attracted growing interest, with approximately 4.5% of individuals with sleep disorders reportedly turning to complementary or alternative medicine [6]. Saffron (*Crocus sativus*) has emerged as a promising candidate due to its bioactive compounds, particularly safranal and crocin, which have demonstrated potential in modulating neurotransmitter activity, reducing oxidative stress, and improving mood and sleep quality [7].

Previous clinical studies support the potential of saffron supplementation to enhance sleep quality and reduce stress. In one randomized controlled trial involving 63 healthy adults aged 18–70 with self-reported sleep problems, saffron extract (14 mg twice daily) significantly improved sleep quality within one week, as measured by the Insomnia Severity Index (ISI), the Restorative Sleep Questionnaire (RSQ), and the Pittsburgh Sleep Diary (PSD) [8]. A subsequent trial, conducted over 28 days with 120 adults experiencing unsatisfactory sleep, further demonstrated improvements in self-reported sleep quality and mood, as assessed by the Pittsburgh Sleep Diary, the Insomnia Symptom Questionnaire (ISQ), the Profile of Mood States, and the Functional Outcomes of Sleep Questionnaire. Additionally, another study explored the acute effects of saffron and its main volatile compound, safranal, under stress conditions, showing that saffron intake delayed peak salivary cortisol and cortisone levels during a laboratory stress test [9]. These effects are likely mediated by the impact of saffron on serotonin reuptake, GABA receptor modulation, and its anti-inflammatory, antioxidant, neuroprotective, and HPA-axis-regulating properties [10].

Despite these promising findings, most of the existing trials have been limited by small sample sizes, which restricts the generalizability of their results. To address this gap, this randomized, double-blind, placebo-controlled study represents a large, nationwide effort aimed at evaluating the effects of saffron extract at two dosages (20 mg and 30 mg) on insomnia symptoms, sleep quality, perceived stress, anxiety and depressive symptoms, and quality of life in adults with moderate insomnia, using a combination of validated self-reported questionnaires.

## 2. Methods

This clinical trial followed a three-arm, parallel-group, double-blind, placebo-controlled design spanning four weeks. The study adhered to the CONSORT guidelines and received ethical approval from the Ethics Committee of the Medical Association of Lower Saxony (Hannover, Germany). The protocol was developed in line with the principles of Good Clinical Practice (GCP) and the Declaration of Helsinki. Trial registration was completed with the German Clinical Trials Register (DRKS) (DRKS00033435). Data collection for eligibility screening, informed consent, participation tracking, and all questionnaire-based outcome assessments was conducted using LimeSurvey Community Edition (version 3.28.5+220405), a secure, web-based platform hosted on a university server at Leibniz University Hannover.

### 2.1 Participants

Participants were recruited nationwide in Germany through press release, radio advertisements and social media campaigns. The recruitment period took place in February and March 2023. Interested individuals were directed to an online screening tool that included the Regensburg Insomnia Scale (RIS), as well as demographic and health-related questions to assess eligibility. Enrollment occurred only after participants had met all eligibility criteria, read the study information, and provided informed consent via a secure online participation form integrated into the screening platform.

#### 2.1.1 Inclusion criteria

Adults aged between 18 and 65 years with moderate insomnia, as indicated by a Regensburg Insomnia Scale (RIS) score greater than 12, were eligible to participate. The RIS is a validated self-report questionnaire used to assess cognitive, emotional, and behavioral symptoms associated with insomnia over the past four weeks [11]. Other criteria included a BMI between 20 and 35 kg/m² and a consistent bedtime routine between 9 pm and midnight.

People with stable thyroid disease (e.g. Hashimoto’s disease, hyperthyroidism or hypothyroidism) or hypertension were included if their medication had not changed for at least two months before the study. Participants were asked to refrain from taking any dietary supplements or medications that affect sleep for two weeks prior to the intervention.

#### 2.1.2 Exclusion criteria

Participants were excluded if they were pregnant, breastfeeding, working rotating or night shifts. Other exclusion criteria were: diagnosed sleep apnea or any chronic sleep disorder for more than one year, serious chronic diseases such as diabetes, cardiovascular disease, active cancer, or neurological disorders, regular use of medications such as antidepressants, antihistamines, beta-blockers, anticoagulants, or psychotropic drugs.

### 2.2. Intervention

The study consisted of a four-week intervention phase during which participants consumed one capsule of a full spectrum standardized saffron extract (either 20 mg or 30 mg, Safr’Inside™, Activ’Inside, Beychac et Caillau, France) or a placebo (maltodextrine) each evening, approximately 60 minutes before bedtime. This saffron extract obtained through the patent EP3490575B1-WO2017EP69200 contained crocins >3%, safranal >0.2%, picrocrocin derivatives >1% and kaempferol derivatives >0.1%, analyzed using the U-HPLC method.

Participants were instructed to maintain their usual dietary habits, physical activity levels, and daily routines throughout the intervention. Adherence was monitored weekly through online questionnaires and confirmed by capsule counts at the end of the study. Participants in the placebo group were provided with a 4 week-bottle of saffron extract capsules as compensation after unblinding.

### 2.3 Outcome measures

#### 2.3.1 Athens Insomnia Scale (AIS) – primary outcome

The AIS is an eight-item self-assessment questionnaire designed to evaluate the severity of insomnia symptoms based on ICD-10 diagnostic criteria [12]. It assesses sleep induction, nighttime and early morning awakenings, total sleep duration, and daytime performance and fatigue. Each item is scored from 0 to 3, with total scores ranging from 0 to 24. Higher scores indicate greater insomnia severity.

#### 2.3.2 Single-item Sleep Quality Scale (SQS)

The SQS measures overall sleep quality over the past seven days using a single visual analog scale, ranging from 0 (terrible) to 10 (excellent) [13]. Participants rated their sleep quality based on their experiences during this period. The question prompts participants to consider various factors, such as sleep duration, ease of falling asleep, frequency of waking during the night, and overall refreshment upon waking.

#### 2.3.3 Epworth Sleepiness Scale (ESS)

The ESS assesses daytime sleepiness by evaluating the likelihood of dozing off in eight common scenarios [14]. Each scenario is rated on a scale from 0 (no likelihood) to 3 (high likelihood), with total scores ranging from 0 to 24. Higher scores indicate increased daytime sleepiness.

#### 2.3.4 Positive and Negative Affect Schedule (PANAS)

The PANAS is a 20-item questionnaire designed to measure positive and negative emotional states [15]. Participants rated items on a five-point Likert scale, reflecting the frequency of specific emotions over the past week. Higher scores indicate stronger emotional responses in the respective domains. Total scores range from 10 to 50 for both positive and negative affect, with higher scores reflecting greater emotional intensity.

#### 2.3.5 Perceived Stress Scale (PSS)

The PSS measures stress levels over the past four weeks using 10 items rated on a five-point scale [16]. Total scores range from 0 to 40, with 0–13 indicating low stress, 14–26 moderate stress, and 27+ high stress.

#### 2.3.6 Patient Health Questionnaire-4 (PHQ-4)

The PHQ-4 is a brief screening tool for symptoms of anxiety and depression [17]. It includes four items rated on a four-point Likert scale, with two items each dedicated to anxiety and depression. Total scores range from 0 to 12, with higher scores suggesting greater symptom severity.

#### 2.3.7 WHO Quality of Life-BREF (WHOQOL-BREF)

The WHOQOL-BREF is derived from the WHOQOL-100 and is suitable for studies where quality of life is not the primary outcome [18]. It assesses quality of life across four domains: physical health, psychological health, social relationships, and the environment. These domains are evaluated using a 5-point Likert interval scale, reflecting the quality of life over the past two weeks [19].

### 2.4 Sample Size Calculation

The sample size estimation was based on detecting an expected effect size of f = 0.25 in AIS scores, using a power of 80% and an alpha level of 5%. This effect size was selected based on previous studies investigating the impact of interventions on sleep quality in adults with moderate insomnia [20]. To account for an anticipated dropout rate of 20%, the final target sample size was set at 150 participants to ensure sufficient power to detect a clinically significant effect.

### 2.5 Randomization and Blinding

Participants were randomly assigned to one of three groups (20 mg saffron extract, 30 mg saffron extract, or placebo) through a block-based randomization process. Randomization was stratified according to age, gender, and baseline RIS scores to ensure balanced group allocation. Both participants and study investigators remained blinded to group assignments until the intervention phase and final data review were complete.

### 2.6 Statistical Methods

Primary analyses were conducted on an intention-to-treat (ITT) basis, including all randomized participants with post-baseline data, and supplemented by per-protocol (PP) analyses to assess robustness of the results. Helmert contrasts were systematically applied for all parameters to compare the combined intervention groups (20 mg and 30 mg saffron) with the placebo group to ensure consistency in the assessment of group effects.

Group differences in the Athens Insomnia Scale (AIS) were analyzed using analysis of covariance (ANCOVA), adjusted for age, sex, body weight, and baseline AIS score. These covariates were preselected based on theoretical and methodological considerations. Post hoc pairwise comparisons of estimated marginal means were performed using Tukey’s method, with p-values adjusted for multiple comparisons. Effect sizes for AIS were calculated using Cohen’s d, based on pooled residual standard deviations from the ANCOVA model.

For the Perceived Stress Scale (PSS), the Kruskal-Wallis test was used due to the non-normal data distribution. Dunn’s test with Holm’s adjustment for multiple comparisons was used for post hoc pairwise comparisons, and effect sizes were calculated using the rank-biserial correlation coefficient (*r*) derived from the Wilcoxon rank sum test.

As a sensitivity analysis, multiple imputations were performed to account for missing ITT data for the AIS and PSS, which were based on change scores calculated from two time points (baseline and endpoint). Missing values were imputed using predictive mean matching (PMM) with five imputed records. Rubin’s rules were used to pool estimates, variances, and test statistics across imputations. For the AIS, ANCOVA was performed directly on the imputed records, while for the PSS, Kruskal-Wallis and Dunn’s tests were applied separately to each imputed record, and the results were pooled to produce the final estimates.

All other parameters were analyzed using linear mixed models (LMMs) since endpoints were collected at least 3 times during the period. The LMMs included fixed effects for time, group, their interaction, and covariates (age, occupation, weight, and baseline scores), as well as random intercepts to account for repeated measures within participants. The most appropriate model structure was selected using the Single-Item Sleep Quality Scale (SQS) as the reference model, and models were fit using the Akaike Information Criterion (AIC). Post hoc group comparisons at specific time points were performed using Tukey-adjusted pairwise tests, while simple slope analysis was used to assess group effects over the entire study period. Beta coefficients (*β*) were derived to quantify effect sizes for comparisons based on the LMM.

All statistical analyses were performed using R software (version 4.4.2) with the following packages: car, emmeans, rstatix, coin, lme4, lmerTest, mice, and dplyr. Statistical significance was set at *P* < .05.

## 3. Results

### 3.1 Study population

A total of 1,768 individuals were assessed for eligibility, of whom 165 participants were randomized into three groups: saffron 30 mg (n = 55), saffron 20 mg (n = 55), and placebo (n = 55) (Figure 1). Of the randomized participants, 158 completed the baseline survey and were included in the intention-to-treat (ITT) analysis, which included all participants who provided at least baseline data. The placebo group included 49 participants, while the saffron 20 mg and saffron 30 mg groups included 55 and 54 participants, respectively. The per-protocol (PP) analysis included 147 participants who adhered strictly to the study protocol.

**Figure 1.**
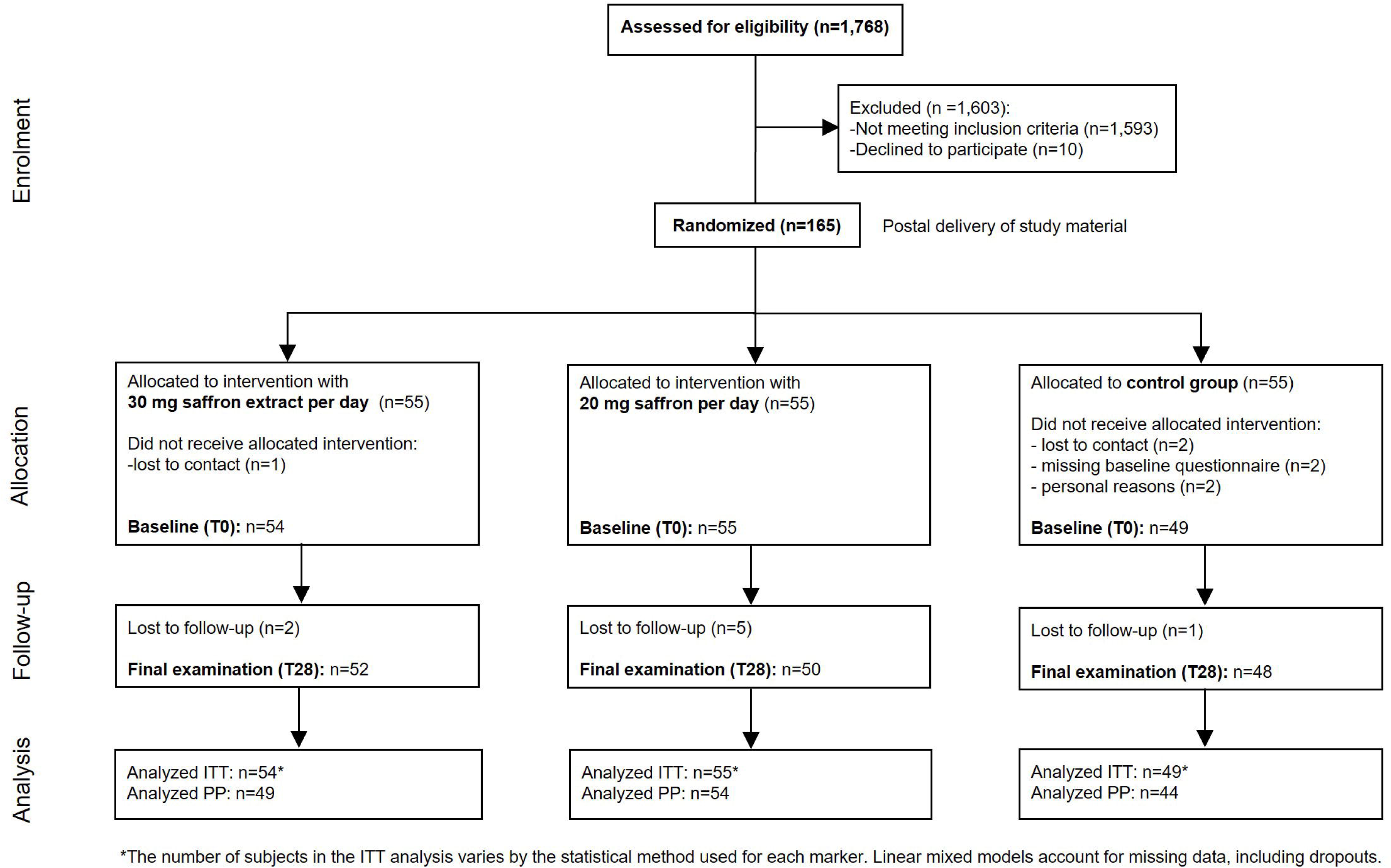
Participant flow diagram according to CONSORT guidelines. The diagram illustrates participant progression through the stages of enrollment, randomization, allocation, follow-up, and analysis.

Reasons for exclusion from the PP population included low adherence to capsule intake (i.e., consuming fewer than 80% of the provided capsules or missing intake for more than seven days), use of certain medications or supplements (e.g., magnesium) for more than three consecutive days, and deviations from predefined sleep and lifestyle criteria. Specifically, participants were excluded if they consumed more than three alcoholic drinks per day, drank caffeinated beverages within four hours before bedtime, or had a bedtime later than 2 a.m. on multiple occasions for social reasons. In the main analysis, exclusion from the PP population also applied if participants had deviations in their sleep diary on three or more occasions or went to bed later than 6 a.m. more than once.

Reasons for non-completion included failure to establish contact (n = 3), not completing the baseline questionnaire on time (n = 2), and unspecified reasons (n = 3), which were grouped under ‘lost contact’ or ‘failure to provide required data.’

Baseline demographic characteristics, including age, BMI, and questionnaire scores, were comparable across groups, with no significant differences detected (Table 2). The participants were mainly women (77%), had a mean age of 44.5 years (SD: 11.5), and had a BMI within normal range.

**Table 1.** Outcome parameters and time of survey.

| Parameter | Baseline<br>T0 | Survey<br>T7 | Survey<br>T14 | Survey<br>T21 | Survey<br>T28 |
| --- | --- | --- | --- | --- | --- |
| **Primary** |  |  |  |  |  |
| AIS | X |  |  |  | X |
| **Secondary** |  |  |  |  |  |
| PANAS | X | X | X | X | X |
| ESS | X | X | X | X | X |
| PSS | X |  |  |  | X |
| SQS | X | X | X | X | X |
| WHOQOL-BREF | X |  | X |  | X |
| PHQ-4 | X |  | X |  | X |

AIS = Athens Insomnia Scale, ESS = Epworth Sleepiness Scale, NSD = non-specific sleep diary, PANAS = Positive and Negative Affect Schedule, PHQ-4 = Patient Health Questionnaire-4, PSS = Perceived Stress Scale, RIS = Regensburg Insomnia Score, SQS = Single-item Sleep Quality Scale, WHOQOL = World Health Organization Quality of Life

**Table 2.**
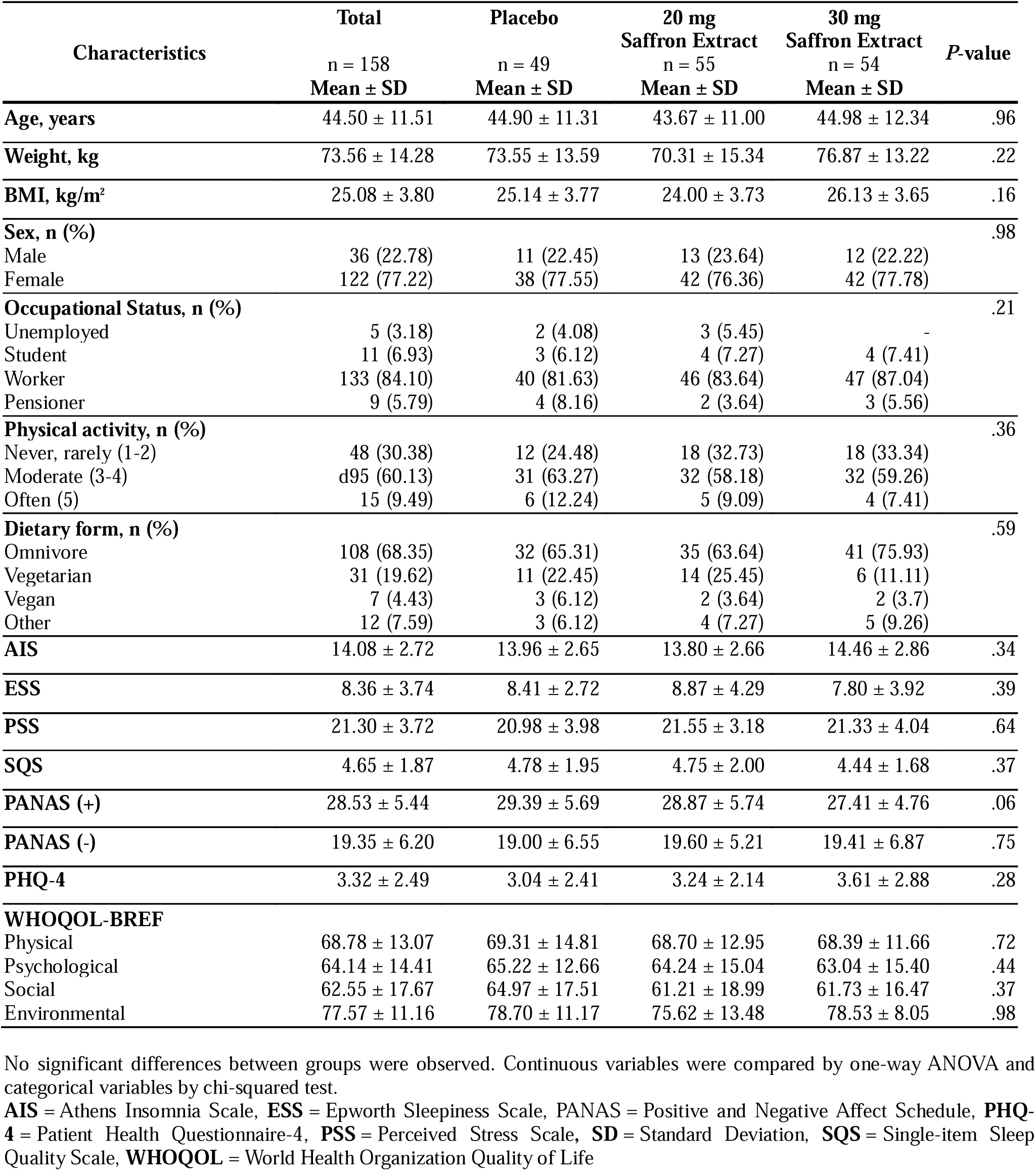
Baseline demographic characteristics of participants.

At baseline, the average Athens Insomnia Scale (AIS) score was 14.08 (SD: 2.72), indicating moderate to severe insomnia symptoms, well above the normal range (0-5). The single-item Sleep Quality Scale (SQS) score was 4.65 (SD: 1.87), indicating suboptimal sleep quality. The Epworth Sleepiness Scale (ESS) mean score of 8.36 (SD: 3.74) is slightly elevated, but still within the threshold for normal daytime sleepiness (< 10).

Stress and mood measures also indicate a moderately distressed population. The Perceived Stress Scale (PSS) mean score of 21.30 (SD: 3.72) falls within the range of moderate stress (14-26). The Positive Affect Schedule (PANAS+) score of 28.53 (SD: 5.44) is slightly below typical normative values (∼30), while the Negative Affect Schedule (PANAS-) score of 19.55

(SD: 6.20) is slightly above average, indicating a slightly more negative emotional state. The Patient Health Questionnaire-4 (PHQ-4) mean score of 3.32 (SD: 2.49) is in the mild range for anxiety and depression symptoms.

Overall, these baseline characteristics suggest that the study population has moderate sleep disturbance, increased stress, and slightly elevated negative affect, which is consistent with the intended target population of individuals experiencing sleep disturbance but not severe psychiatric illness.

### 3.2 Primary endpoint: Athens Insomnia Scale (AIS)

In the ITT analysis, a significant group effect was observed for the AIS scores (*P* = .035), indicating differences between the intervention and placebo groups. Post hoc comparisons revealed that the combined saffron groups (20 mg and 30 mg) showed a statistically significant reduction in insomnia severity compared to placebo, as determined by Helmert contrasts (*P* = .027, d = 0.41).

On average, AIS scores decreased by −1.96 points (SD = 3.14) in the saffron 30 mg group, - 1.54 points (SD = 2.24) in the saffron 20 mg group, and −0.71 points (SD = 2.27) in the placebo group (Table 3). Analysis of individual AIS components showed that the most pronounced differences between the saffron and placebo groups were observed in sleep induction (30 mg saffron extract: −0.314, 20 mg saffron extract: −0.223) and sleep duration (30 mg saffron extract: −0.255, 20 mg saffron extract: −0.323) (data not shown). The PP analysis yielded consistent results, with slightly larger reductions in AIS scores observed in the saffron groups, particularly in the 30 mg group (−2.45 points, SD = 3.20), while the placebo group exhibited only a modest improvement (−0.89 points, SD = 2.37) (data not shown). Figure 2A illustrates the changes in AIS scores over time, adjusted for baseline AIS values and relevant covariates, depicting reductions across groups. As a sensitivity analysis within the ITT population, multiple imputation was performed to address missing AIS data at T28, and the results were consistent with those from the complete case analysis (data not shown).

**Figure 2.**
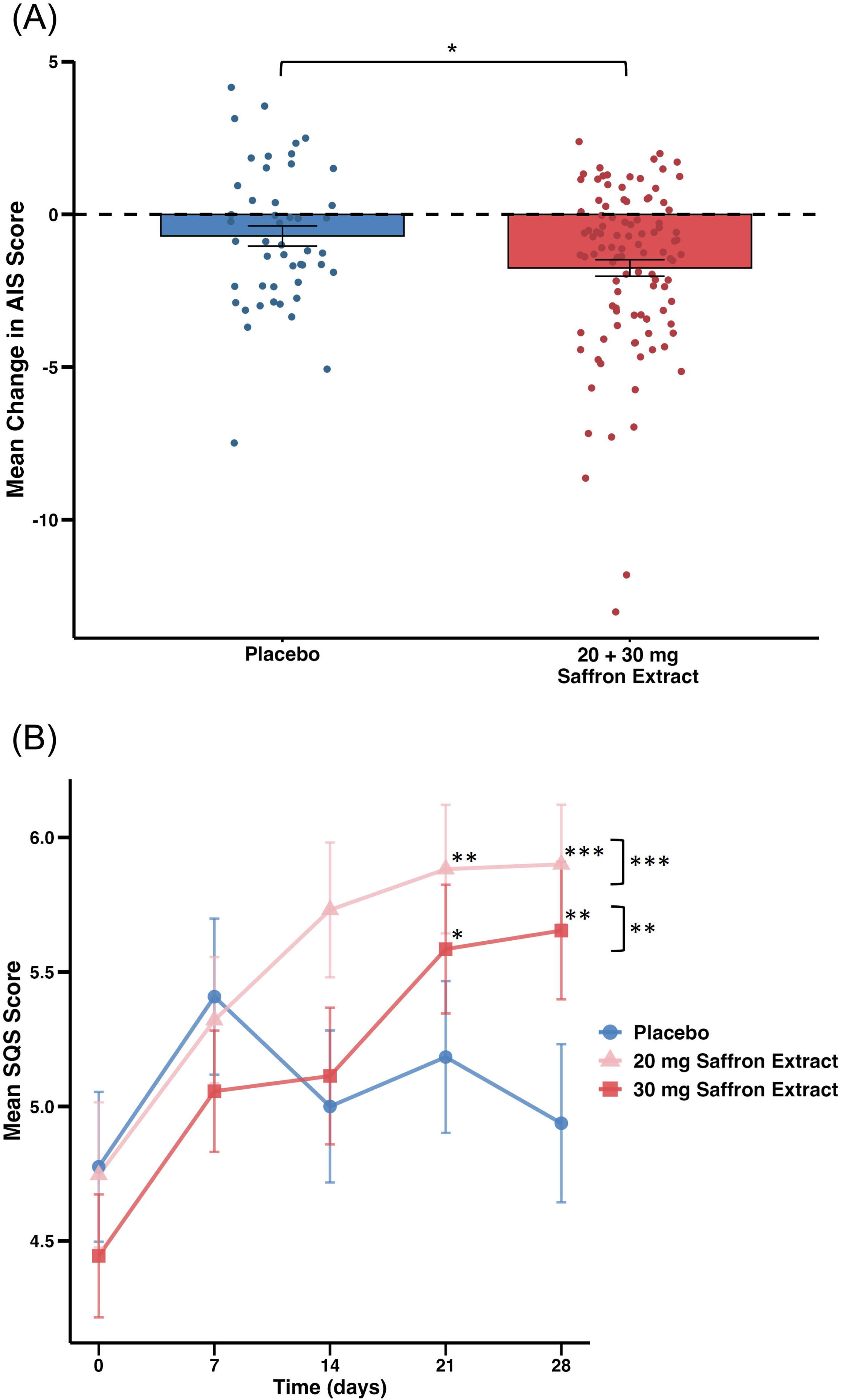
Effect of saffron extract on insomnia symptoms and self-reported sleep quality compared with placebo. **(A) Mean change in Athens Insomnia Scale (AIS) scores.** Scatter points represent individual participant AIS score changes from baseline (T0) to day 28 (T28) in the placebo group (n = 48) and saffron intervention group (n = 102; 20 mg, n = 50; 30 mg, n = 52; combined for analysis). Bars represent group means ± standard error of the mean (SEM). The horizontal dashed line indicates no change in AIS score (mean change = 0). Statistical analysis was conducted using an analysis of covariance (ANCOVA) within an intention-to-treat (ITT) framework, adjusted for age, sex, body weight, and AIS baseline score. Helmert contrasts were used to compare mean AIS score changes. *Asterisks indicate statistically significant differences (*P* < .05). **(B) Mean scores of the single-item sleep quality scale (SQS).** Scores from baseline (T0) to day 28 (T28) in the placebo group (n = 49), 20 mg saffron extract group (n = 55), and 30 mg saffron extract group (n = 54) are shown. Scores represent observed means ± SEM. Statistical analysis was conducted using a linear mixed model (LMM) within an ITT framework, adjusted for age, occupation, and baseline SQS scores. Pairwise comparisons were conducted with Tukey adjustments. Asterisks at specific time points indicate significant post hoc pairwise comparisons, while those to the right of brackets represent overall group differences over time (simple slope analysis), with P values Tukey-adjusted (\**P* < .05, \*\**P* < .01, \*\*\**P* < .001).

**Table 3.**
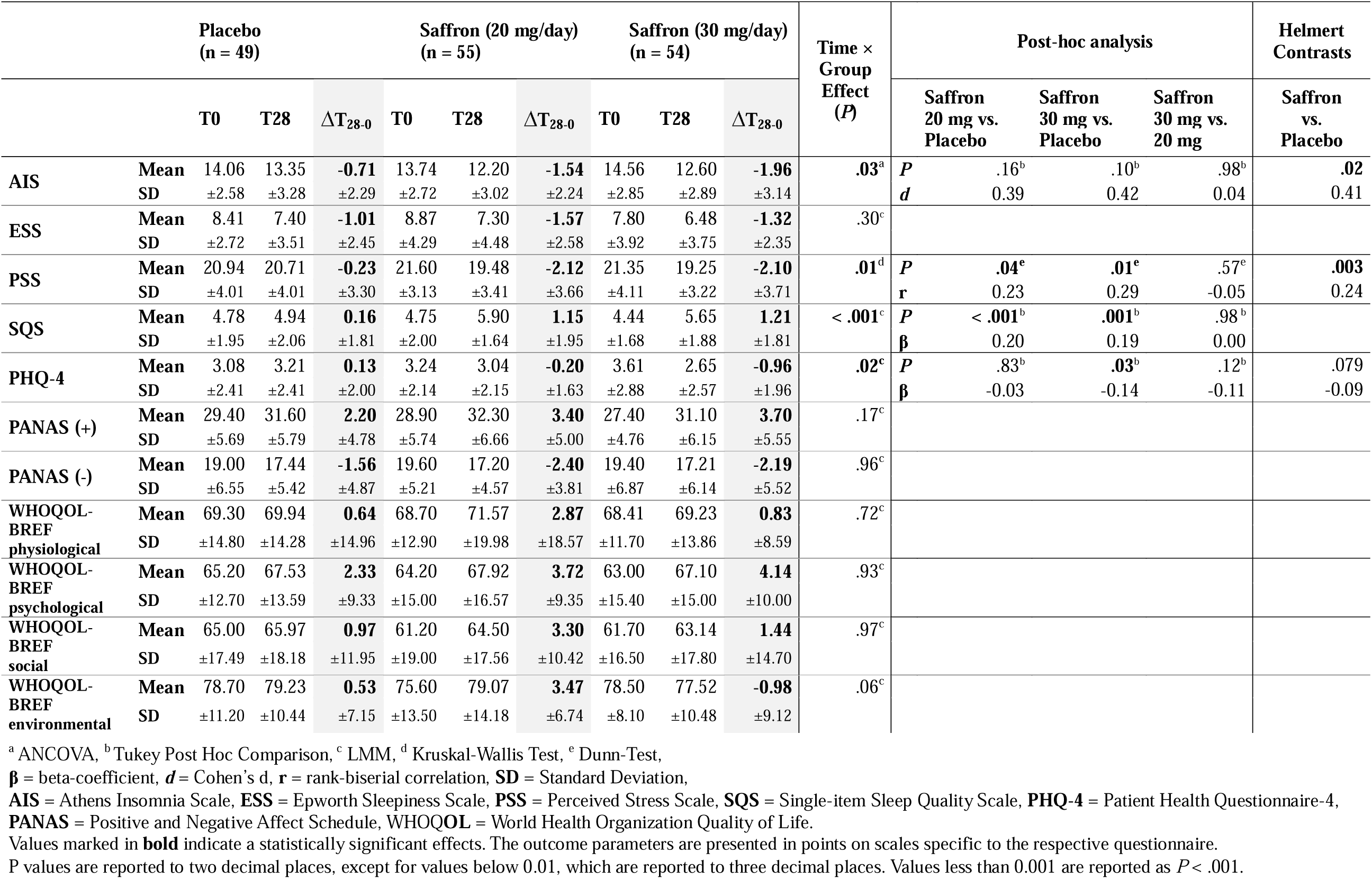
Change in self-reported questionnaire scores from baseline to week 4 (T28) in the intention-to-treat (ITT) analysis.

### 3.3 Secondary endpoints

Weekly change in sleep quality, as assessed by the Single-Item Sleep Quality Scale (SQS), improved significantly in both saffron intervention groups compared to placebo. In the ITT analysis, saffron 20 mg and 30 mg were associated with improved perceived sleep quality over 4 weeks (*P* < .001 and *P* = .001, respectively), as determined by linear mixed model (LMM) simple slope analysis. Differences between the saffron and placebo groups became statistically significant after three weeks of supplementation and remained significant thereafter (Figure 2B). The PP analysis also yielded significant results (*P* < .01), with effect sizes similar as in the ITT analysis (data not shown).

Perceived stress levels, measured by PSS, showed significant reductions in both saffron groups compared to placebo in the ITT analysis (Figure 3). The 30 mg saffron group demonstrated a mean reduction of −2.10 points (SD = ±3.71, *P* = .012), while the 20 mg group showed a comparable improvement of −2.12 points (SD = ±3.66, *P* = .043). The placebo group showed no significant changes with a mean reduction of −0.23 points (SD = ±3.30, *P* = .578). The PP analysis showed similar results, with significant differences between the groups. Analysis of the Epworth Sleepiness Scale revealed no significant differences in daytime sleepiness in either the ITT or PP analyses (Table 3). To assess the robustness of the findings, multiple imputation was applied to the ITT dataset to account for missing PSS data at T28, yielding results that aligned closely with the complete case analysis (data not shown).

**Figure 3.**
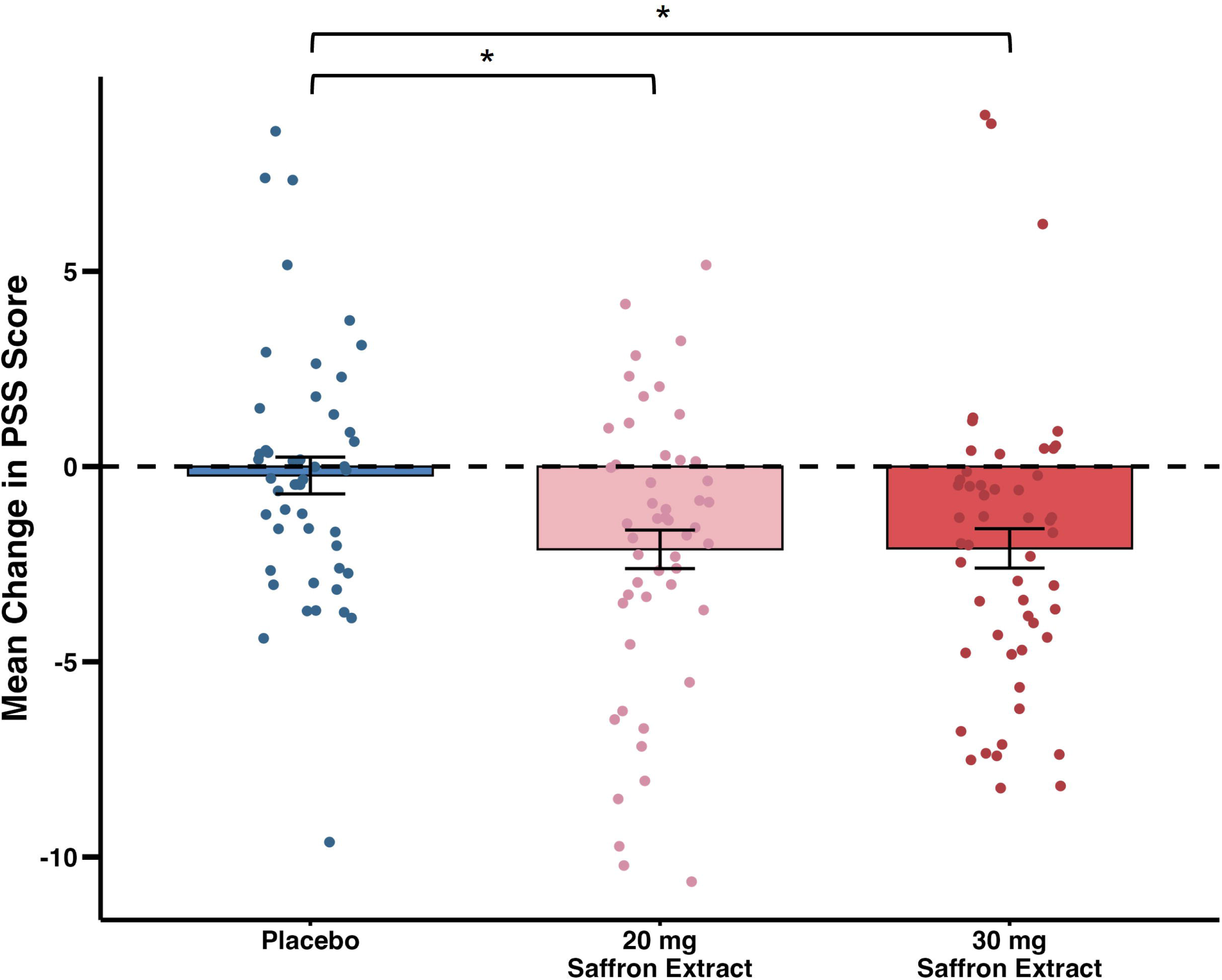
Change in Perceived Stress Scale (PSS) Scores from Baseline to Day 28. Scatter points represent individual participant PSS score changes in the placebo group (n = 48), 20 mg saffron extract group (n = 55), and 30 mg saffron extract group (n = 54); points are jittered slightly for better visibility. Bars represent changes in group means ± standard error of the mean (SEM). The horizontal dashed line indicates no change in PSS score (mean change = 0). Statistical analysis was conducted using the Kruskal-Wallis test due to non-normal data distribution. Pairwise comparisons were performed using Dunn’s test, and the analysis followed an intention-to-treat (ITT) approach. *Asterisks indicate statistically significant between-group differences (*P* < .05) compared with the placebo group.

The Patient Health Questionnaire-4 (PHQ-4), a measure of depression and anxiety symptoms, showed a significant reduction in the 30 mg saffron group compared to placebo (*P* = .034, β = −0.14), while the 20 mg saffron group exhibited no meaningful change (*P* = .837, β = −0.03) in the ITT analysis (Table 3). The mean change in the PHQ-4 score was −0.96 (SD = ±1.96) for the 30 mg saffron group, −0.20 (SD = ±1.63) for the 20 mg group, and 0.13 (SD = ±2.00) for the placebo group. Figure 4 illustrates the progression of PHQ-4 scores across the intervention period, showing minimal changes in the placebo and 20 mg saffron groups, with the 30 mg group showing a more noticeable reduction.

**Figure 4.**
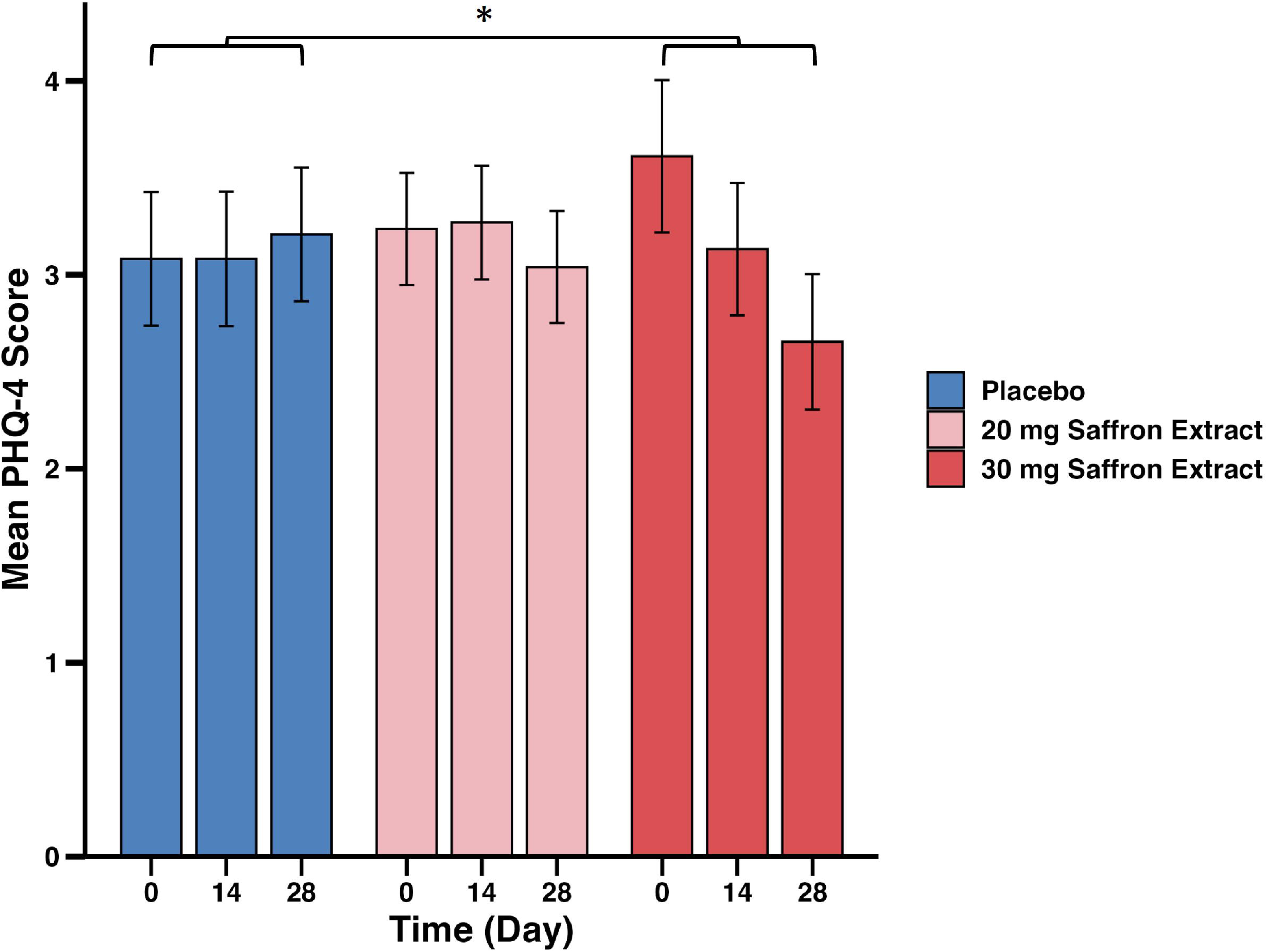
Mean Patient Health Questionnaire-4 (PHQ-4) scores from baseline to day 28. Scores are shown for the placebo group (n = 49), 20 mg saffron extract group (n = 55), and 30 mg saffron extract group (n = 54). Values represent observed means ± standard error of the mean (SEM). Statistical analysis was conducted using a linear mixed model (LMM) within an intention-to-treat (ITT) framework, adjusted for age, occupation, and baseline PHQ-4 scores. Asterisk indicates a statistically significant between-group difference in change scores (*P* < .05) based on Tukey-adjusted pairwise comparisons.

### 3.4 Adverse events

Saffron supplementation was well tolerated across groups. Gastrointestinal discomfort was the most frequently reported adverse event, occurring in 10 cases in the 30 mg saffron group, 6 cases in the 20 mg group, and 4 cases in the placebo group throughout the 4-week trial (Table 4). Other adverse events, including headaches and fatigue, were rare and did not lead to study discontinuation. No serious adverse events were reported, and adherence remained high across all groups (Figure 1). Dropout rates were similar across groups, with overall retention remaining robust.

**Table 4.**
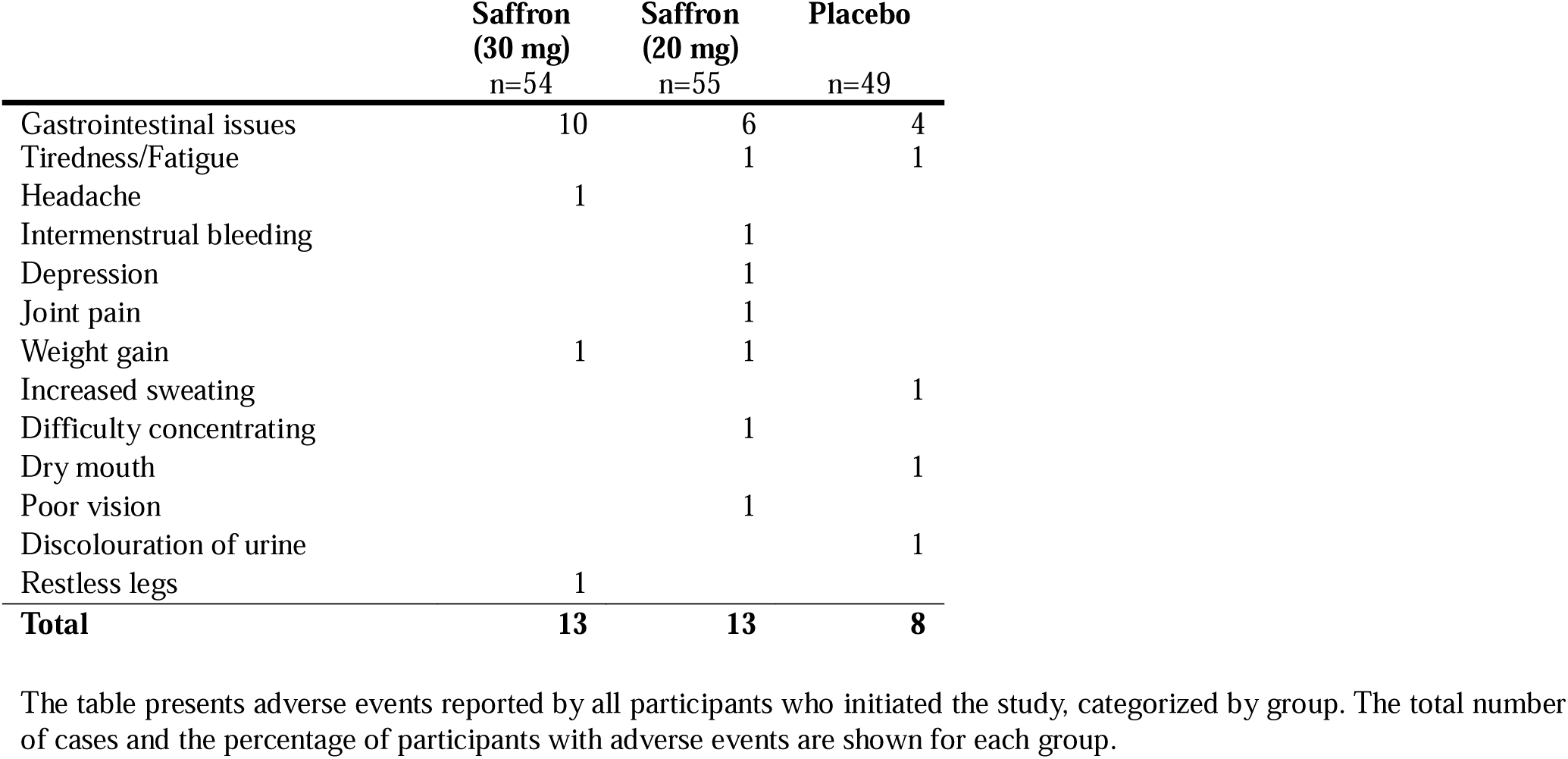
Self-reported adverse events.

|  | **Saffron**<br>**(30 mg)**<br>**n=54** | **Saffron**<br>**(20 mg)**<br>**n=55** | **Placebo**<br>**n=49** |
| --- | --- | --- | --- |
| Gastrointestinal issues | 10 | 6 | 4 |
| Tiredness/Fatigue |  | 1 | 1 |
| Headache | 1 |  |  |
| Intermenstrual bleeding |  | 1 |  |
| Depression |  | 1 |  |
| Joint pain |  | 1 |  |
| Weight gain | 1 | 1 |  |
| Increased sweating |  |  | 1 |
| Difficulty concentrating |  | 1 |  |
| Dry mouth |  |  | 1 |
| Poor vision |  | 1 |  |
| Discolouration of urine |  |  | 1 |
| Restless legs | 1 |  |  |
| **Total** | **13** | **13** | **8** |

The table presents adverse events reported by all participants who initiated the study, categorized by group. The total number of cases and the percentage of participants with adverse events are shown for each group.

## 4. Discussion

### 4.1 The effect of saffron on sleep, stress and mood

To our knowledge, this is the largest randomized controlled trial to date evaluating the effects of saffron extract supplementation on insomnia severity, sleep quality, and psychological well-being in adults reporting moderate insomnia. Our findings indicate that saffron extract supplementation can significantly improve these outcomes. Specifically, participants receiving saffron extract showed measurable improvements in insomnia severity (AIS), self-reported sleep quality (SQS), perceived stress (PSS), and psychological distress (PHQ-4) compared to placebo. Although previous studies have hinted at saffron’s potential benefits for sleep and stress reduction, many of these trials were limited by small sample sizes, short durations, or a lack of rigorous control groups. In our double-blind, placebo-controlled study, 165 participants were randomized (150 completers: saffron 30 mg: 52; saffron 20 mg: 50; placebo: 48), addressing these gaps and providing robust support for the potential of both 20 mg and 30 mg saffron extract as non-pharmacological interventions to enhance sleep and alleviate stress-related outcomes.

The Athens Insomnia Scale (AIS) scores showed a statistically significant reduction when combining the saffron groups (30 mg and 20 mg) versus placebo (*P* = .035, d = 0.45), indicating a small to moderate effect size. However, separate comparisons for saffron 20 mg and 30 mg versus placebo (*P* =0.161 and *P* = .109) were not significant after adjusting for multiple comparisons. These findings are consistent with prior studies demonstrating saffron’s efficacy for improving sleep, which also reported small but statistically significant effects on insomnia severity [8]. The Single-Item Sleep Quality Scale (SQS), with its weekly assessments, demonstrated that all groups experienced an initial improvement in sleep quality within the first seven days. However, while the placebo group showed a temporary increase, its effect diminished over time, whereas the saffron supplementation groups continued to improve, reaching statistical significance after three weeks. This pattern suggests that the early improvements observed across all groups may include a placebo response, but the sustained benefits in the saffron groups likely reflect a true treatment effect. These findings align with prior research highlighting the rapid onset of action of saffron in improving self-reported sleep outcomes [8,20,21].

Daytime sleepiness, assessed by the Epworth Sleepiness Scale (ESS), remained unaffected across groups. This is consistent with previous findings [21] suggesting that saffron’s impact on daytime fatigue may be limited to specific populations with higher baseline fatigue levels. The absence of significant changes in daytime sleepiness in the present study could be attributed to low baseline ESS scores, which limited the potential for meaningful improvement.

Stress levels, as measured by the Perceived Stress Scale (PSS), significantly decreased in both saffron groups compared to placebo. Notably, this is the first time that a 20 mg dose of saffron extract produced a stress-reducing effect comparable to the 30 mg dose. This may be due to a higher baseline stress level in this study compared to previous research, or a higher safranal content in the saffron extract used here, which is approximately 10 times higher than in extracts used in earlier studies [8,22]. Previous studies have shown that saffron supplementation can modulate both psychological and biological stress responses, including delaying peak salivary cortisol levels and dampening acute stress perceptions in response to controlled stressors, potentially through modulation of the hypothalamic-pituitary-adrenal (HPA) axis [9]. However, findings on saffron’s effects on cortisol remain mixed [20], as another trial reported no significant impact on evening cortisol levels, suggesting that its stress-reducing effects may be mediated by other mechanisms, such as modulation of neurotransmitters or melatonin regulation. Although administered as a single dose, saffron extract has been found to attenuate stress-induced physiological changes such as reductions in heart rate variability in healthy adults [23].

The 30 mg dose of saffron extract was also associated with significant improvements in overall scores on the Patient Health Questionnaire-4 (PHQ-4), particularly due to reductions in the depression subscore. Saffron’s effects on psychological well-being may be attributed to its influence on multiple neurotransmitter systems. Active compounds such as safranal, crocin, and crocetin have been shown to inhibit the reuptake of serotonin and dopamine while also exhibiting N-methyl-D-aspartate (NMDA) receptor antagonism and gamma-aminobutyric acid (GABA)-A agonism, mechanisms implicated in mood regulation and stress resilience [7]. A recent meta-analysis found that saffron’s effects on depressive and anxiety symptoms were comparable to those of selective serotonin reuptake inhibitors (SSRIs), but with fewer adverse events [24]. While the precise pathways remain under investigation, reductions in psychological distress may also contribute to improved sleep quality, as elevated depressive symptoms are a known risk factor for insomnia [25].

Saffron’s effects on sleep may not only be attributed to its influence on neurotransmitter systems but also to its antioxidant and anti-inflammatory properties. Its bioactive compounds help inhibit nuclear factor kappa B (NFκB) activation, leading to reduced oxidative stress and lower expression of pro-inflammatory cytokines such as tumor necrosis factor-alpha (TNF-α) and interleukin-6 (IL-6), while simultaneously upregulating antioxidant enzymes like superoxide dismutase and catalase [26]. Since chronic inflammation is linked to sleep disturbances, these effects may contribute to improving sleep quality, particularly in individuals with inflammation-related sleep disorders. Additionally, pro-inflammatory cytokines regulate key circadian genes, including Clock and BMAL1, which are essential for maintaining a stable sleep-wake cycle [27]. By modulating these pathways, saffron may help stabilize circadian rhythms, reduce daytime fatigue, and promote overall sleep quality.

Furthermore, these regulatory effects may enhance melatonin production, which plays a crucial role in sleep onset and duration [28].

### 4.2 Strengths and limitations

One of the major strengths of this study is its larger sample size compared to existing research on saffron and sleep [29]. The use of validated online questionnaires allowed for nationwide participation in Germany, which facilitated ease of participation and adherence, contributing to a very low dropout rate. The study design using a randomized, double-blind, placebo-controlled methodology enhances internal validity and minimizes bias. In addition, the inclusion of two saffron dosage groups allowed for a dose-response analysis, providing further insight into the potential therapeutic effects of saffron.

A notable limitation of this study is its reliance on self-reported sleep measures, which may introduce bias and limit objectivity in the assessment of sleep outcomes. Future studies should incorporate more reliable actigraphy devices or polysomnography to improve the accuracy of objective sleep measures.

As expected with the dose allowed in nutritional supplements, the observed effect sizes were in the small to medium range. This highlights the need to combine saffron supplementation with other lifestyle interventions, such as improving sleep hygiene by reducing screen time at bedtime and limiting sedentary behavior, or cognitive behavioral therapy for insomnia, to achieve meaningful benefits. Further research is needed to identify potential high responders—individuals who may derive greater benefits from saffron—by considering factors such as baseline sleep quality, genetic predispositions, or interactions with other dietary and behavioral factors.

Furthermore, the lack of physiological markers such as cortisol or melatonin limits the ability to assess saffron’s modulation of circadian rhythms. Inclusion of these biomarkers, as done in previous studies, or evaluation of other relevant markers could help elucidate the underlying mechanisms driving the sleep and stress-related effects of saffron. However, it was technically unfeasible to include these markers in the clinical design due to the nationwide recruitment of participants and the instability of melatonin at room temperature after postal delivery, as observed by validation tests. Finally, it should be noted that the predominantly female study population (77%) may limit the generalizability of the findings. Although women are generally more affected by insomnia than men, future studies should aim to include a more balanced representation of the sexes to ensure broader applicability of the results.

### 4.3 Conclusions

As a nationwide home-based trial, this randomized, double-blind, placebo-controlled clinical study contributes to the growing evidence that Safr’inside ^TM^ supplementation (20 or 30 mg for 4 weeks) may improve sleep symptoms, stress, and psychological well-being in individuals with moderate levels of insomnia and stress. Significant improvements were observed for insomnia severity (AIS), self-reported sleep quality (SQS), stress (PSS), and psychological distress (PHQ-4), but effects on daytime sleepiness (ESS), mood (PANAS), and quality of life (WHOQOL-BREF) were not significant.

These findings suggest that saffron supplementation may serve as a well-tolerated, adjunctive intervention alongside other evidence-based lifestyle strategies to improve insomnia symptoms. Future research should focus on identifying individuals most likely to benefit from saffron supplementation, including those with clinically diagnosed insomnia, and further investigating its potential impact on sleep and stress outcomes. Longer intervention periods, more detailed sleep diary assessments, and the inclusion of reliable objective biomarkers may help to refine this understanding and clarify potential effects beyond self-reported experiences.

## Data Availability

All data produced in the present study are available upon reasonable request to the authors.

## Acknowledgements

The authors thank the Activ’Inside team (Beychac et Caillau, France) for their collaboration in designing the study. Data collection and analysis were conducted independently by the academic team at the Institute of Food and One Health, Leibniz University Hannover. Manuscript preparation was led by the academic team, with editorial feedback provided by the sponsor.

This study was financially supported by Activ’Inside following initial contact from the authors.

## Author contributions

**Julius Schuster*:** Conceptualization, Methodology, Validation, Formal Analysis, Investigation, Data Curation, Writing – Original Draft, Writing – Review & Editing, Visualization, Supervision, Project Administration

**Christin Mundhenke*:** Methodology, Formal Analysis, Investigation, Data Curation, Writing – Original Draft, Visualization

**Hannah Nordsieck:** Investigation, Data Curation

**Camille Pouchieu:** Conceptualization, Methodology, Writing – Review & Editing

**Line Pourtau:** Conceptualization, Methodology, Writing – Review & Editing

**Andreas Hahn:** Conceptualization, Supervision, Resources, Funding Acquisition, Writing – Review & Editing

**\***both authors contributed equally

## Clinical Trial Registration

**Trial name:** Effect of a saffron extract on sleep parameters in healthy adults with poor sleep: a randomized, double blind, 3 arm, placebo-controlled clinical study.

**URL:** https://drks.de/search/en/trial/DRKS00033435

**DRKS-ID:** DRKS00033435

## Declaration of Generative AI and AI-Assisted Technologies in the Writing Process

During the preparation of this work, the authors used DeepL and ChatGPT in order to assist with language phrasing and translation accuracy. After using these tools, the authors reviewed and edited the content as needed and take full responsibility for the content of the publication.

## Abbreviations

AIS: Athens Insomnia Scale
ANCOVA: Analysis of Covariance
ANOVA: Analysis of Variance
BMAL1: Brain and Muscle Arnt-Like Protein 1
ESS: Epworth Sleepiness Scale
GABA: Gamma-aminobutyric acid
HPA: Hypothalamic-Pituitary-Adrenal
HRV: Heart Rate Variability
ITT: Intention-To-Treat
LMM: Linear Mixed Model
NREM: Non-Rapid-Eye-Movement
PANAS: Positive and Negative Affect Schedule
PHQ-4: Patient Health Questionnaire 4
PP: Per-Protocol
PSS: Perceived Stress Scale
REM: Rapid-Eye-Movement
RIS: Regensburg Insomnia Scale
ROS: Reactive Oxygen Species
SQS: Single-item Sleep Quality Scale
WHOQOL-BREF: World Health Organization Quality of Life-BREF

